# Mitral regurgitation trajectories after transcatheter aortic valve replacement are phenotype specific across low-flow aortic stenosis subtypes

**DOI:** 10.64898/2026.08.07.26359941

**Authors:** Akarsh Sharma, Esha Vaish, Eileen Galvani, Annapoorna S. Kini, Samin K. Sharma, Stamatios Lerakis

## Abstract

**Objectives:** Mitral regurgitation (MR) evolution after transcatheter aortic valve replacement (TAVR) in low-flow aortic stenosis (LFAS) is poorly characterized. We evaluated MR trajectories across LFAS phenotypes, predictors of MR worsening, and associations with clinical outcomes.

**Methods:** We retrospectively studied 614 LFAS patients undergoing TAVR: low-flow high-gradient (LFHG; n=153, 24.9%), classical low-flow low-gradient (cLFLG; n=155, 25.2%), and paradoxical low-flow low-gradient (pLFLG; n=306, 49.8%). MR severity was abstracted from clinical echocardiography reports using a 6-level ordinal scale. MR worsening was defined as a ≥1-grade increase from baseline MR at ∼30 days or ∼1 year. Multivariable logistic models identified predictors of MR worsening. Kaplan–Meier and Cox models evaluated associations of MR trajectory and LFAS subtype with all-cause death, heart failure hospitalization (HFH), and their composite.

**Results:** Among 614 LFAS patients, 443 had 30-day and 290 had 1-year echocardiographic follow up. At 30 days, MR trajectory differed significantly across LFAS phenotypes, with the highest rate of worsening in cLFLG and the lowest in LFHG. At 1 year, unadjusted MR trajectory distributions did not differ significantly across phenotypes. In adjusted logistic models, cLFLG remained independently associated with MR worsening at both timepoints. MR worsening was associated with worse unadjusted outcomes at 30 days but was not independently associated with the composite endpoint after multivariable adjustment. LFAS phenotype, particularly cLFLG, remained the dominant predictor of adverse clinical outcomes.

**Conclusions:** MR evolution after TAVR is phenotype-specific: cLFLG patients have the highest risk of MR worsening and lowest event-free survival, supporting phenotype-informed post-TAVR surveillance.

## Introduction

Transcatheter aortic valve replacement (TAVR) has revolutionized the management of severe aortic stenosis (AS), particularly in patients deemed high or prohibitive surgical risk.^1^ However, the heterogeneity of AS phenotypes presents unique challenges in predicting post-procedural outcomes and optimizing patient selection. Low-flow aortic stenosis (LFAS), characterized by a stroke volume index <35 mL/m², encompasses distinct subtypes with varying pathophysiological mechanisms and prognoses.^2^

The three primary LFAS phenotypes, low-flow high gradient (LFHG), classical low-flow, low-gradient (cLFLG), and paradoxical low-flow, low-gradient (pLFLG)—differ fundamentally in their underlying hemodynamics.^2–3^ LFHG typically represents true severe AS with preserved contractility, cLFLG is characterized by reduced left ventricular ejection fraction (LVEF <50%) suggesting myocardial dysfunction, while pLFLG paradoxically combines preserved LVEF with low-flow, often due to pronounced concentric remodeling and small cavity size.^4^ These phenotypic distinctions have proven prognostically important, with cLFLG patients experiencing particularly poor outcomes even after successful TAVR.^5^

Concomitant mitral regurgitation (MR) is prevalent in LFAS patients, affecting 30-50% of those undergoing TAVR.^6–7^ The complex interplay between AS and MR creates a challenging clinical scenario, as relief of AS may theoretically improve MR through reduced left ventricular pressures and improved hemodynamics. However, the trajectory of MR following TAVR in this population is not fully understood nor clearly predictable with competing mechanisms of potential improvement as well as progression.

Previous studies have demonstrated conflicting results regarding MR evolution post-TAVR, with improvement rates ranging from 20-60% and worsening observed in 10-25% of patients.^7^ These disparate findings likely reflect the heterogeneous nature of the studied populations and the failure to account for distinct AS phenotypes. The prognostic implications of MR changes after TAVR remains particularly controversial, with some studies suggesting increased mortality with persistent or worsening MR, while others find no significant association.^8^

The phenotype-specific behavior of MR after TAVR in LFAS patients has not been systematically evaluated. Understanding these patterns is crucial for several reasons: (1) it may inform pre-procedural risk stratification and patient counseling, (2) it has implications for post-TAVR surveillance protocols, and (3) it may identify subgroups warranting closer surveillance and future study of staged mitral intervention strategies.

This study aimed to characterize the evolution of MR across LFAS phenotypes following TAVR and evaluate its potential association with clinical outcomes (all-cause mortality and heart failure hospitalization). We hypothesized that MR trajectory would vary significantly between LFHG, cLFLG, and pLFLG subtypes due to their distinct pathophysiological substrates, and that these differences would have phenotype-specific prognostic implications.

## Methods

### Study Population

Adults (≥ 18 years) who underwent TAVR at a high-volume quaternary care hospital in New York City between January 2019 and December 2022 were included if they fulfilled hemodynamic LFAS criteria (stroke volume index < 35 mL/m² and aortic valve area ≤1.0 cm²). Reasons for exclusion included prior valve surgery (n=98), concomitant procedures (n=45), in-hospital mortality (n=31), and inadequate echocardiographic assessment (n=58).

Low-flow subtypes were classified as:

- LFHG (Low-Flow High-Gradient): mean gradient ≥ 40 mmHg
- cLFLG (Classical Low-flow, low-gradient): LVEF < 50% and mean gradient < 40 mmHg
- pLFLG (Paradoxical Low-flow, low-gradient): LVEF ≥ 50% and mean gradient < 40 mmHg

A multidisciplinary Heart Team determined patient eligibility for TAVR based on comprehensive evaluation including cardiovascular conditions, functional status assessment using New York Heart Association classification and Kansas City Cardiomyopathy Questionnaire scores, frailty assessment, and procedural risk stratification using the Society of Thoracic Surgeons Predicted Risk of Mortality (STS-PROM) score. The transcatheter valve systems utilized included Edwards SAPIEN 3 (Edwards Lifesciences, Irvine, CA), Medtronic CoreValve, Evolut R, and Evolut PRO (Medtronic, Minneapolis, MN), and Boston Scientific Acurate Neo and Neo2 (Boston Scientific, Marlborough, MA).

Baseline demographic, clinical, and echocardiographic data were extracted retrospectively from the electronic medical record. The study protocol was approved by the Institutional Review Board at the Icahn School of Medicine at Mount Sinai, with waiver of informed consent due to the retrospective design.

### Echocardiography

Transthoracic echocardiographic studies were performed. The mechanism of MR was not recorded. MR severity was abstracted from routine clinical echocardiography reports and mapped to a six-level ordinal scale: 0 none/trace, 1 mild, 2 mild-to-moderate, 3 moderate, 4 moderate-to-severe, and 5 severe. Moderate-or-greater MR was defined as grade ≥3. Quantitative MR parameters were not consistently available.

Post-TAVR echocardiograms were obtained within defined windows: early (15-90 days, targeting 30 days) and late (185-545 days, targeting 365 days). When multiple echocardiograms existed within a window, the study closest to the target day was selected.

MR change from baseline was categorized as:

- Worsened: increase by ≥ 1 grade
- Stable: no change in grade
- Improved: decline by ≥ 1 grade

### Outcomes

The primary clinical outcome was a composite of all-cause mortality or heart failure hospitalization. Secondary clinical outcomes included all-cause mortality and heart failure hospitalization as individual endpoints. The primary echocardiographic outcome was change in MR grade.

For clinical outcome analyses involving MR trajectory, MR worsening was treated as a landmark exposure at the corresponding echocardiographic timepoint. Patients with events before the landmark, or echocardiograms obtained after a composite outcome event, were excluded from the corresponding landmark analysis; follow up for outcomes began at the landmark timepoint.

### Statistical Analysis

Continuous variables are presented as medians and interquartile ranges, with statistical testing performed using analysis of variance (ANOVA) or Kruskal-Wallis tests for non-normally distributed variables. Categorical variables are presented as number (percentage of total group) and compared using χ² test with post-hoc pairwise comparisons when significant differences were detected.

Multivariable logistic regression, using the models described below, was performed to identify predictors of MR worsening (versus stable/improved) at both 30-day and 1-year timepoints. Age and STS score were analyzed as continuous variables (per 1 year and per 1% increase, respectively). Baseline MR grade was modeled as a continuous ordinal variable in the primary multivariable models; moderate-or-greater MR, defined as grade ≥3, was used for descriptive baseline characterization. Complete case analysis was performed, with sample sizes varying by model due to missing data. Exploratory interaction terms between LFAS subtype and MR worsening were tested for clinical outcomes but were not included in final primary models because of limited subgroup event counts and model instability. Statistical significance was defined as p<0.05.

Cox proportional hazards regression was performed to evaluate associations between MR worsening, LFAS phenotype, and clinical outcomes using landmark analyses at the corresponding echocardiographic timepoint. MR worsening was treated as a landmark exposure. Patients with composite events before the landmark, echocardiograms occurring on or after a composite-event date, or missing required model covariates were excluded from the corresponding model. Age and STS-PROM were analyzed as continuous variables. Baseline MR grade was modeled as a continuous ordinal variable on the 1–5 clinical scale.

Three hierarchical models were used where event counts allowed: a basic model adjusted for LFAS phenotype, age, sex, and baseline MR grade; an extended model additionally adjusted for STS-PROM and prosthesis–patient mismatch; and a full model additionally adjusted for chronic kidney disease, atrial fibrillation, and coronary artery disease. Extended and full models were restricted to patients with complete STS-PROM and prosthesis–patient mismatch data. For 1-year secondary outcomes, extended and full models were suppressed when events-per-variable was critically low, or estimates were unstable.

The basic model included fundamental adjustments selected a priori for clinical relevance and parsimony: LFAS phenotype, age, sex, and baseline MR grade. Age was modeled continuously per 1-year increase, and baseline MR grade was modeled as a continuous ordinal variable on the 1–5 clinical scale. The extended model included procedure-related factors and risk assessment variables: the basic model plus STS-PROM, modeled continuously per 1% increase, and prosthesis–patient mismatch. STS-PROM has demonstrated predictive value for mortality in TAVR patients, and prosthesis–patient mismatch has been associated with incomplete MR resolution after aortic valve replacement.^9,10^ The full model takes the extended model one step further by incorporating additional comorbidities that have clinically been demonstrated to impact MR significantly: chronic kidney disease^11^, atrial fibrillation^12^, and clinically-significant coronary artery disease^13^. Clinically significant coronary artery disease was defined as one or more of the following: history of myocardial infarction, percutaneous coronary intervention, and/or coronary artery bypass grafting.

Interaction terms between LFAS subtype and MR worsening were tested but excluded from final models when limited subgroup events were detected. Analyses were performed separately for composite outcome (death or HF hospitalization), all-cause mortality, and HF hospitalization. Kaplan-Meier curves were generated for the overall cohort stratified by MR trajectory and LFAS subtypes. Log-rank tests compared survival curves with post-hoc pairwise comparisons when indicated.

Statistical analyses were performed using R (version 4.3.0) in RStudio. Two-sided p < 0.05 defined statistical significance. Pairwise comparisons of MR trajectory distributions were adjusted using the Holm method.

## Results

### Study population

A total of 614 patients met inclusion criteria. LFAS phenotype distribution was LFHG 153 (24.9%), cLFLG 155 (25.2%), and pLFLG 306 (49.8%).

### Baseline characteristics by subtype

Baseline characteristics differed across LFAS subtypes (Table 1). Median age was slightly lower in LFHG 81 (74–86) years compared with 83 for pLFLG (77-88) and 83 (76–89) years for cLFLG (p = 0.023). Male sex was least prevalent in pLFLG (54%) compared to cLFLG (80%) and LFHG (61%) (p<0.001).

**Table 1.** Baseline characteristics by low-flow aortic stenosis phenotype.

| Characteristic | LFHG<br>N = 153 | cLFLG<br>N = 155 | pLFLG<br>N = 306 | p-value<sup>1</sup> |
| --- | --- | --- | --- | --- |
| Age, Median (Q1, Q3) | 81 (74, 86) | 83 (76, 89) | 83 (77, 88) | **0.023** |
| Male, n (%) | 93 (61%) | 124 (80%) | 165 (54%) | **<0.001** |
| STS-PROM, Median (Q1, Q3) | 3.1 (1.9, 5.0) | 3.9 (2.7, 5.9) | 3.2 (2.1, 5.0) | **<0.001** |
| Chronic kidney disease, n (%) | 43 (28%) | 71 (46%) | 99 (32%) | **0.002** |
| Atrial fibrillation, n (%) | 45 (29%) | 71 (46%) | 108 (35%) | **0.010** |
| Coronary artery disease, n (%) | 68 (44%) | 107 (69%) | 133 (43%) | **<0.001** |
| **Prosthesis–patient mismatch, n (%)** |  |  |  | 0.16 |
| No | 73 (82%) | 70 (79%) | 136 (88%) |  |
| Yes | 16 (18%) | 19 (21%) | 19 (12%) |  |
| **Baseline MR grade, n (%)** |  |  |  | **0.015** |
| 1 | 49 (32%) | 35 (23%) | 109 (36%) |  |
| 2 | 81 (53%) | 84 (54%) | 157 (51%) |  |
| 3 | 11 (7.2%) | 27 (17%) | 28 (9.2%) |  |
| 4 | 9 (5.9%) | 5 (3.2%) | 7 (2.3%) |  |
| 5 | 3 (2.0%) | 4 (2.6%) | 5 (1.6%) |  |
| Moderate-or-greater MR, n (%) | 23 (15%) | 36 (23%) | 40 (13%) | **0.018** |

<sup>1</sup> Kruskal-Wallis rank sum test; Pearson's Chi-squared test. Data presented as median (Q1, Q3) or n (%). Bold p-values indicate p < 0.05.

Compared with pLFLG and LFHG, cLFLG patients had a higher prevalence of chronic kidney disease (46% vs 32% and 28%), atrial fibrillation (46% vs 35% and 29%), and coronary artery disease (69% vs 43% and 44%) (all p≤0.01). Baseline moderate-or-greater MR (grade ≥3) was also more frequent in cLFLG (23%) than in pLFLG (13%) or LFHG (15%) (p=0.018). Rates of prosthesis–patient mismatch did not differ significantly between subtypes (p=0.16).

### MR trajectories after TAVR Early trajectory (30-day)

Among 443 patients with baseline and 30-day echocardiographic follow up, 67 (15.1%) experienced MR worsening from baseline (Table 2; Figure 1). The overall distribution of MR change (improved / stable / worsened) differed significantly across LFAS phenotypes (χ²=16.88, df=4, p=0.002) with higher rates of early MR worsening in cLFLG (26.2%) than in pLFLG (13.0%) or LFHG (9.1%) (Table 2a). Post-hoc pairwise comparisons (Table 2b) revealed that MR-change distribution in cLFLG differed from both pLFLG (Holm-adjusted p=0.008) and LFHG (Holm-adjusted p=0.006), whereas pLFLG and LFHG did not differ (Holm-adjusted p=0.44).

**Figure 1.**
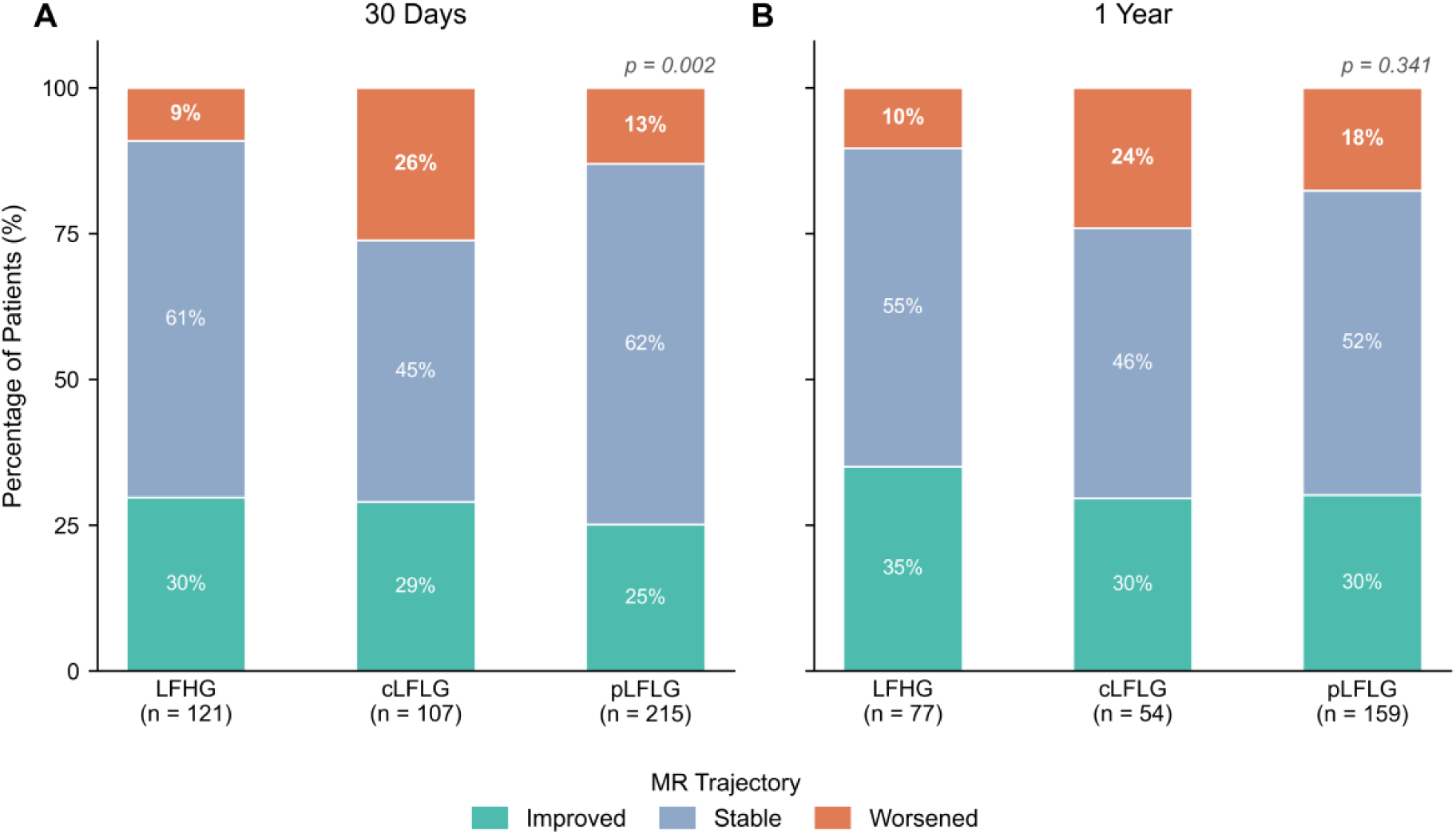
Mitral regurgitation trajectory after TAVR by low-flow aortic stenosis phenotype. Stacked bars show the proportion of patients with improved, stable, or worsened MR at 30 days and 1 year after TAVR. MR worsening was defined as a ≥1-grade increase from baseline; improvement was defined as a ≥1-grade decrease. At 30 days, MR trajectory differed significantly across LFAS phenotypes (p = 0.002), with the highest rate of worsening in cLFLG. At 1 year, the unadjusted MR trajectory distribution did not differ significantly across phenotypes (p = 0.341).

**Table 2.** Mitral regurgitation trajectory after TAVR by low-flow aortic stenosis phenotype.

**Panel A. MR change distribution by LFAS phenotype**
| Timepoint | MR Change | LFHG (N = 121) | cLFLG (N = 107) | pLFLG (N = 215) | p-value |
| --- | --- | --- | --- | --- | --- |
| 30 days | Improved | 36 (29.8%) | 31 (29.0%) | 54 (25.1%) | **0.002** |
| 30 days | Stable | 74 (61.2%) | 48 (44.9%) | 133 (61.9%) |  |
| 30 days | **Worsened** | **11 (9.1%)** | **28 (26.2%)** | **28 (13.0%)** |  |
| 1 year | Improved | 27 (35.1%) | 16 (29.6%) | 48 (30.2%) | 0.341 |
| 1 year | Stable | 42 (54.5%) | 25 (46.3%) | 83 (52.2%) |  |
| 1 year | Worsened | 8 (10.4%) | 13 (24.1%) | 28 (17.6%) |  |

**Panel B. Pairwise comparisons of MR change distribution (Holm correction)**
| Timepoint | Comparison | $\chi^2$ | df | Raw p-value | Holm-adjusted p-value |
| --- | --- | --- | --- | --- | --- |
| 30 days | **LFHG vs cLFLG** | **12.512** | **2** | **0.002** | **0.006** |
| 30 days | LFHG vs pLFLG | 1.659 | 2 | 0.436 | 0.436 |
| 30 days | **cLFLG vs pLFLG** | **11.174** | **2** | **0.004** | **0.007** |
| 1 year | LFHG vs cLFLG | 4.416 | 2 | 0.110 | 0.330 |
| 1 year | LFHG vs pLFLG | 2.215 | 2 | 0.330 | 0.661 |
| 1 year | cLFLG vs pLFLG | 1.156 | 2 | 0.561 | 0.661 |

Data presented as n (column %). Percentages represent the proportion of patients within each LFAS subtype at the given timepoint.
p-values in Panel A are from overall chi-square tests comparing the full Improved/Stable/Worsened distribution simultaneously across all three LFAS phenotypes (df = 4).
p-values in Panel B are from pairwise chi-square tests (df = 2) with Holm correction for multiple comparisons. Bold values indicate $p < 0.05$ .
MR worsening defined as $\geq 1$ -grade increase from baseline; improvement defined as $\geq 1$ -grade decrease.
Moderate-or-greater MR = grade $\geq 3$ .
Abbreviations: cLFLG, classical low-flow low-gradient aortic stenosis; LFHG, low-flow high-gradient aortic stenosis; LFAS, low-flow aortic stenosis; MR, mitral regurgitation; pLFLG, paradoxical low-flow low-gradient aortic stenosis; TAVR, transcatheter aortic valve replacement.

### Late trajectory (1-year)

Among 290 patients with 1-year echocardiograms, 49 (16.9%) had worsening of MR compared with baseline (Table 2, Figure 1). In contrast to the early timepoint, the distribution of MR change at 1 year did not differ significantly across LFAS phenotypes (χ²=4.51, df=4, p=0.34), and no pairwise comparison between subtypes remained significant after multiplicity adjustment (all Holm-adjusted p≥0.33).

### Predictors of MR worsening Early MR worsening (30-day)

In multivariable logistic regression at 30 days with LFHG as the reference group (Table 3a), the cLFLG phenotype was independently associated with higher odds of MR worsening across all models (full model OR 4.93, 95% CI 1.79–15.4; all p≤0.003). In contrast, pLFLG did not differ significantly from LFHG in any model (full model OR 1.81, 95% CI 0.70–5.30; p=0.24). Higher baseline MR grade was associated with lower odds of early MR worsening (full model OR 0.24 per grade increase, 95% CI 0.13–0.44; p<0.001). Female sex was associated with lower odds of early MR worsening in the basic model (OR 0.47, 95% CI 0.24–0.89; p=0.025), although this association was attenuated and no longer significant after additional adjustment. In the fully adjusted model, chronic kidney disease emerged as an independent risk factor for 30-day MR worsening (OR 2.78, 95% CI 1.28–6.12; p=0.010). Age, STS score, prosthesis–patient mismatch, atrial fibrillation, and coronary artery disease were not significant predictors in the full model.

**Table 3.**
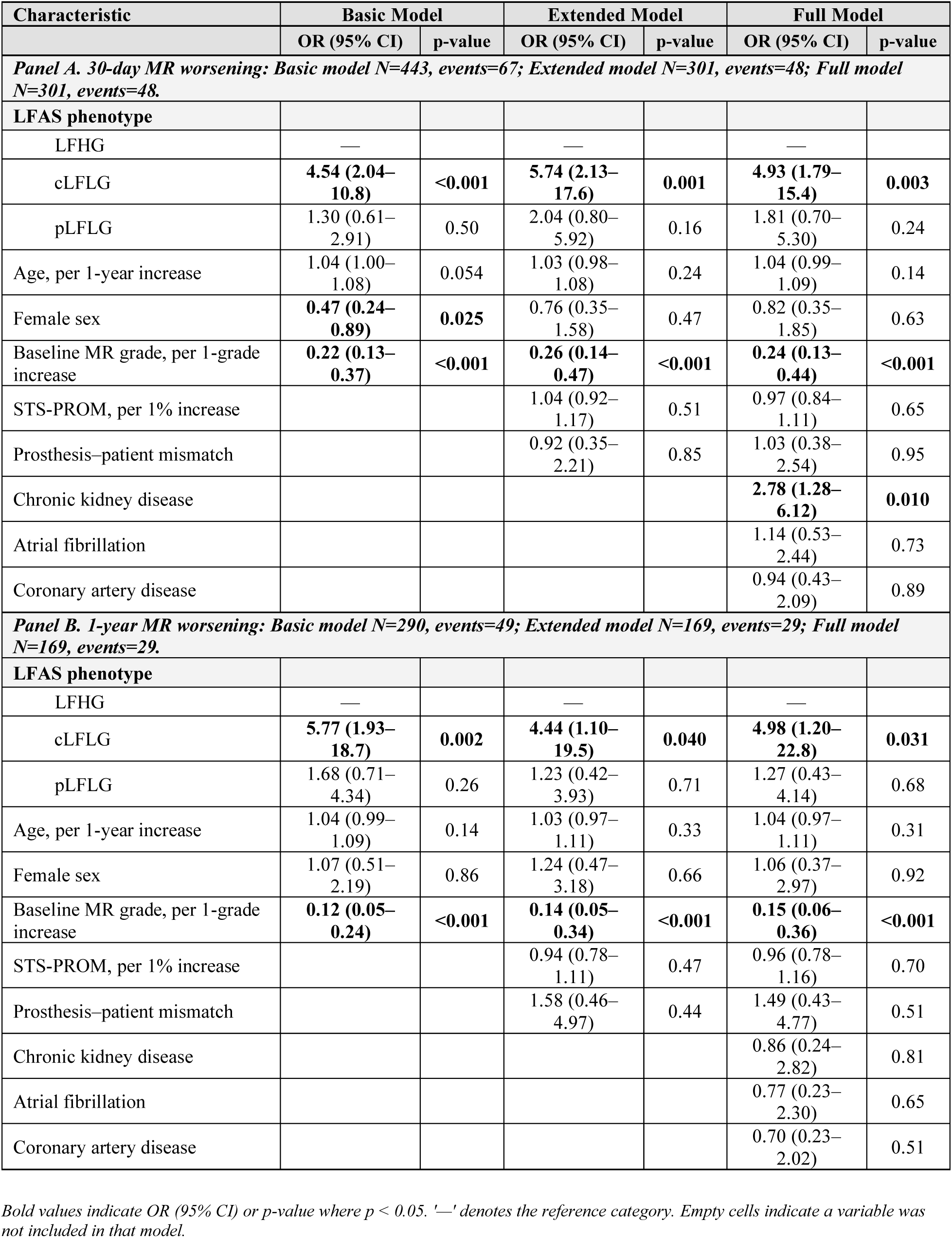
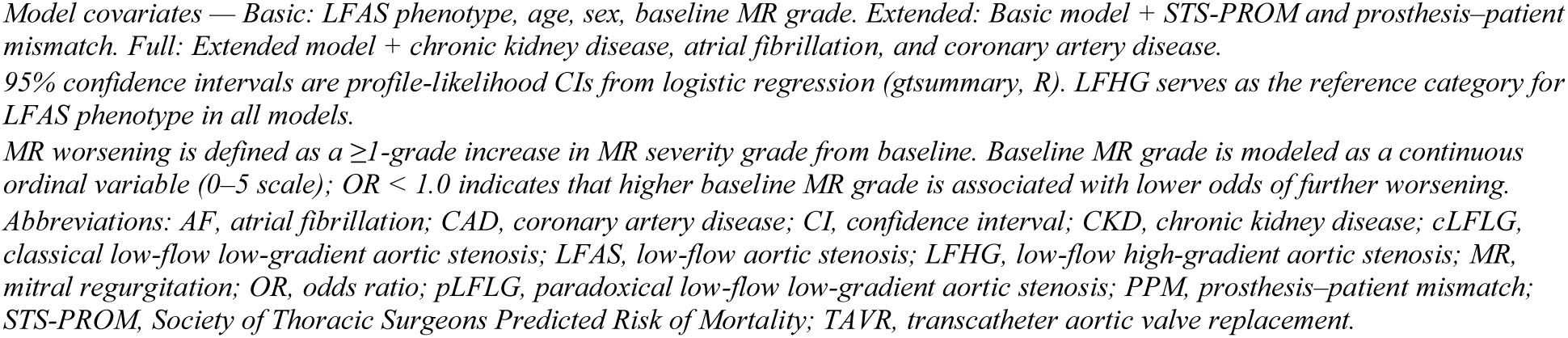
Logistic regression models for mitral regurgitation worsening after TAVR.

### Late MR worsening (1-year)

In multivariable logistic regression at 1 year (Table 3b) with LFHG as the reference group, the cLFLG phenotype remained independently associated with higher odds of MR worsening across all models (full model OR 4.98, 95% CI 1.20–22.8; p=0.031). In contrast, pLFLG did not differ significantly from LFHG in any model (full model OR 1.27, 95% CI 0.43–4.14; p = 0.68). Although the unadjusted 1-year MR-change distribution did not differ significantly across LFAS phenotypes, cLFLG remained independently associated with MR worsening in adjusted binary logistic models.

Baseline MR grade remained inversely associated with late MR worsening (full model OR 0.15 per grade increase, 95% CI 0.06–0.36; p<0.001).

We next examined associations between LFAS phenotype, MR worsening, and clinical outcomes.

### Clinical outcomes Outcomes by LFAS subtype

Clinical outcomes varied by phenotype (Table 4). Compared with LFHG and pLFLG, cLFLG had higher rates of all-cause mortality (34% vs 11% and 16%), heart failure hospitalization (19% vs 7% and 9%), and the composite endpoint of death or heart failure hospitalization (45% vs 17% and 22%) during follow up (all p<0.001). Median follow up also differed across phenotypes, with shorter follow up in cLFLG and pLFLG compared with LFHG.

**Table 4.** Clinical outcomes by low-flow aortic stenosis phenotype.

| Outcome | LFHG<br>(N = 153) | cLFLG<br>(N = 155) | pLFLG<br>(N = 306) | p-value |
| --- | --- | --- | --- | --- |
| All-cause mortality, n (%) | 17 (11%) | 52 (34%) | 48 (16%) | **<0.001** |
| Heart failure hospitalization, n (%) | 10 (7%) | 29 (19%) | 27 (9%) | **<0.001** |
| Composite death or HFH, n (%) | 26 (17%) | 70 (45%) | 66 (22%) | **<0.001** |
| Median follow up, months (IQR) | 27.7 (12.5–<br>37.2) | 15.5 (5.9–<br>26.5) | 17.8 (11.9–<br>27.1) | **<0.001** |

Abbreviations: LFHG, low-flow high-gradient; cLFLG, classical low-flow low-gradient; pLFLG, paradoxical low-flow low-gradient; HFH, heart failure hospitalization; IQR, interquartile range. Composite outcome defined as all-cause death or first heart failure hospitalization, whichever occurred first. Categorical outcomes compared using the chi-square test; follow up time compared using the Kruskal–Wallis test. Bold p-values indicate statistical significance ( $p < 0.05$ ).

### Landmark Cox models for composite outcomes

Among 443 patients in the 30-day landmark cohort, 98 (22.1%) experienced the composite outcome of death or heart failure hospitalization after the landmark timepoint (Table 5). In the basic Cox model, 30-day MR worsening was associated with higher risk of the composite outcome (HR 1.81, 95% CI 1.11–2.96; p=0.017), but this association was attenuated and no longer significant after further adjustment for STS score and comorbidities (full model HR 1.33, 95% CI 0.73–2.44; p=0.358).

**Table 5.** Cox regression models for clinical outcomes after TAVR by MR worsening and LFAS phenotype.

|  |  |  |  | MR Worsening |  | cLFLG vs. LFHG¹ |  | pLFLG vs. LFHG¹ |  |
| --- | --- | --- | --- | --- | --- | --- | --- | --- | --- |
| Outcome | Model | N | Events | HR (95% CI) | p-value | HR (95% CI) | p-value | HR (95% CI) | p-value |
| Panel A. 30-Day Landmark Cohort (N = 443) |  |  |  |  |  |  |  |  |  |
| Composite outcome† | Basic | 443 | 98 | 1.81 (1.11–2.96) | **0.017** | 3.77 (2.07–6.86) | **<0.001** | 1.80 (1.00–3.25) | 0.051 |
|  | Extended | 301 | 68 | 1.47 (0.81–2.65) | 0.206 | 4.25 (2.07–8.75) | **<0.001** | 1.98 (0.96–4.09) | 0.065 |
|  | Full | 301 | 68 | 1.33 (0.73–2.44) | 0.358 | 3.83 (1.83–8.02) | **<0.001** | 1.90 (0.92–3.92) | 0.084 |
| All-cause mortality | Basic | 443 | 69 | 1.45 (0.80–2.65) | 0.220 | 6.09 (2.76–13.4) | **<0.001** | 2.83 (1.27–6.32) | **0.011** |
|  | Extended | 301 | 51 | 1.66 (0.84–3.28) | 0.149 | 4.55 (1.90–10.9) | **<0.001** | 2.52 (1.04–6.14) | **0.042** |
|  | Full‡ | 301 | 51 | 1.59 (0.79–3.21) | 0.197 | 4.11 (1.68–10.1) | **0.002** | 2.50 (1.03–6.09) | **0.044** |
| Heart failure hospitalization | Basic | 443 | 41 | 2.59 (1.25–5.38) | **0.011** | 2.35 (1.00–5.50) | 0.050 | 1.06 (0.46–2.44) | 0.895 |
|  | Extended‡ | 301 | 24 | 1.34 (0.50–3.61) | 0.561 | 4.00 (1.31–12.2) | **0.015** | 1.08 (0.34–3.47) | 0.898 |
|  | Full‡ | 301 | 24 | 1.09 (0.39–3.07) | 0.872 | 3.37 (1.06–10.7) | **0.039** | 1.06 (0.33–3.40) | 0.922 |
| Panel B. 1-Year Landmark Cohort (N = 290) |  |  |  |  |  |  |  |  |  |
| Composite outcome† | Basic | 290 | 36 | 0.92 (0.36–2.34) | 0.861 | 7.51 (2.63–21.4) | **<0.001** | 1.84 (0.66–5.10) | 0.244 |
| All-cause mortality§ | Basic | 290 | 22 | 0.26 (0.05–1.27) | 0.095 | 7.31 (1.88–28.5) | **0.004** | 2.06 (0.56–7.59) | 0.278 |
| Heart failure hospitalization§ | Basic | 290 | 16 | 2.66 (0.85–8.34) | 0.094 | 9.28 (1.86–46.2) | **0.007** | — | — |

† Composite outcome: all-cause death or heart failure hospitalization.
‡ Events-per-variable was low in selected 30-day models; estimates are reported for completeness and should be interpreted with caution.
§ At 1 year, only Basic models are presented because Extended and Full models were restricted to patients with complete STS-PROM and PPM data and had low event counts/unstable estimates. The pLFLG estimate for 1-year heart failure hospitalization was additionally non-estimable because zero HFH events occurred among pLFLG patients with 1-year MR worsening.
<sup>1</sup> LFHG (low-flow high-gradient) is the reference category for all LFAS phenotype comparisons.
**Model definitions:** Basic — adjusted for LFAS phenotype, age, sex, and baseline MR grade (continuous, scale 1–5). Extended — Basic + STS-PROM + prosthesis–patient mismatch. Full — Extended + CKD + atrial fibrillation + CAD. Extended and Full models are restricted to patients with complete STS-PROM and PPM data: N = 301 (30-day landmark), N = 169 (1-year landmark). This explains the N drop from 443 to 301 and from 290 to 169 when moving from Basic to Extended/Full models.
**Note:** Bold p-values indicate $p < 0.05$ . — indicates an estimate that was non-estimable due to complete separation or critically low EPV. All models use LFHG as the reference LFAS phenotype.
**Abbreviations:** AF, atrial fibrillation; CAD, coronary artery disease; CI, confidence interval; CKD, chronic kidney disease; cLFLG, classical low-flow low-gradient aortic stenosis; EPV, events per variable; HFH, heart failure hospitalization; HR, hazard ratio; LFAS, low-flow aortic stenosis; LFHG, low-flow high-gradient aortic stenosis; MR, mitral regurgitation; pLFLG, paradoxical low-flow low-gradient aortic stenosis; PPM, prosthesis–patient mismatch; STS-PROM, Society of Thoracic Surgeons Predicted Risk of Mortality; TAVR, transcatheter aortic valve replacement.

Across all models, LFAS phenotype remained a strong predictor of adverse outcomes. Compared with LFHG, patients with cLFLG had approximately four-fold higher risk of the composite outcome (full model HR 3.83, 95% CI 1.83–8.02; p<0.001), whereas pLFLG showed a non-significant trend toward higher risk (HR 1.90, 95% CI 0.92–3.92; p=0.084). In the fully adjusted model, chronic kidney disease was also independently associated with increased risk (HR 2.11, 95% CI 1.22–3.64; p=0.008), while age showed a modest inverse association with the composite outcome (HR 0.96 per year, 95% CI 0.93–0.99; p=0.014). Baseline MR grade, STS score, prosthesis–patient mismatch, atrial fibrillation, and coronary artery disease were not significant predictors.

Interaction terms between LFAS subtype and 30-day MR worsening were tested in exploratory models and were not statistically significant (all interaction p>0.60).

Among 290 patients in the 1-year landmark cohort, 36 (12.4%) experienced the composite outcome of death or heart failure hospitalization after the landmark timepoint. In basic Cox models using LFHG as the reference phenotype, 1-year MR worsening was not associated with the composite outcome (HR 0.92, 95% CI 0.36–2.34; p=0.861). In contrast, cLFLG remained strongly associated with higher risk of the composite outcome (HR 7.51, 95% CI 2.63–21.4; p<0.001), whereas pLFLG was not significantly associated with risk (HR 1.84, 95% CI 0.66–5.10; p=0.244). Extended and full 1-year models were limited by missing STS-PROM/prosthesis–patient mismatch data and low event counts and were therefore not emphasized.

Exploratory models including interaction terms between LFAS subtype and 1-year MR worsening did not demonstrate significant interactions (all interaction p-values>0.10; data not shown). Kaplan–Meier curves stratified by LFAS subtype alone showed the lowest event-free survival in cLFLG patients (log-rank p≈0.0001).

### Secondary Outcomes

For all-cause mortality in the 30-day landmark cohort, 69 deaths occurred among 443 patients in the basic model. Thirty-day MR worsening was not significantly associated with mortality in the basic model (HR 1.45, 95% CI 0.80–2.65; p=0.22) or full model (HR 1.59, 95% CI 0.79–3.21; p=0.20). In contrast, cLFLG was strongly associated with mortality in both the basic model (HR 6.09, 95% CI 2.76–13.44; p<0.001) and full model (HR 4.11, 95% CI 1.68–10.05; p=0.002). For heart failure hospitalization in the 30-day landmark cohort, 41 events occurred among 443 patients in the basic model. 30-day MR worsening was associated with HF hospitalization in the basic model (HR 2.59, 95% CI 1.25–5.38; p=0.011), but this association was attenuated after full adjustment (HR 1.09, 95% CI 0.39–3.07; p=0.87). cLFLG remained associated with HF hospitalization in the full model (HR 3.37, 95% CI 1.06–10.7; p=0.039).

At the 1-year landmark, 22 deaths and 16 heart failure hospitalizations occurred among 290 patients in the basic models. 1-year MR worsening was not significantly associated with mortality or HF hospitalization, whereas cLFLG remained associated with higher risk of both mortality and HF hospitalization in basic models.

### Kaplan-Meier Survival Analyses

Kaplan–Meier analyses demonstrated worse unadjusted event-free survival among patients with 30-day MR worsening compared with those with stable or improved MR after the 30-day landmark (log-rank p<0.001; Figure 2); however, this association was attenuated and no longer significant after multivariable adjustment. 1-year MR worsening was not associated with a difference in unadjusted event-free survival (log-rank p=0.98). When stratified by LFAS phenotype, event-free survival differed significantly across subtypes in the full LFAS cohort (log-rank p<0.0001; Figure 3), with the lowest event-free survival observed among cLFLG patients.

**Figure 2.**
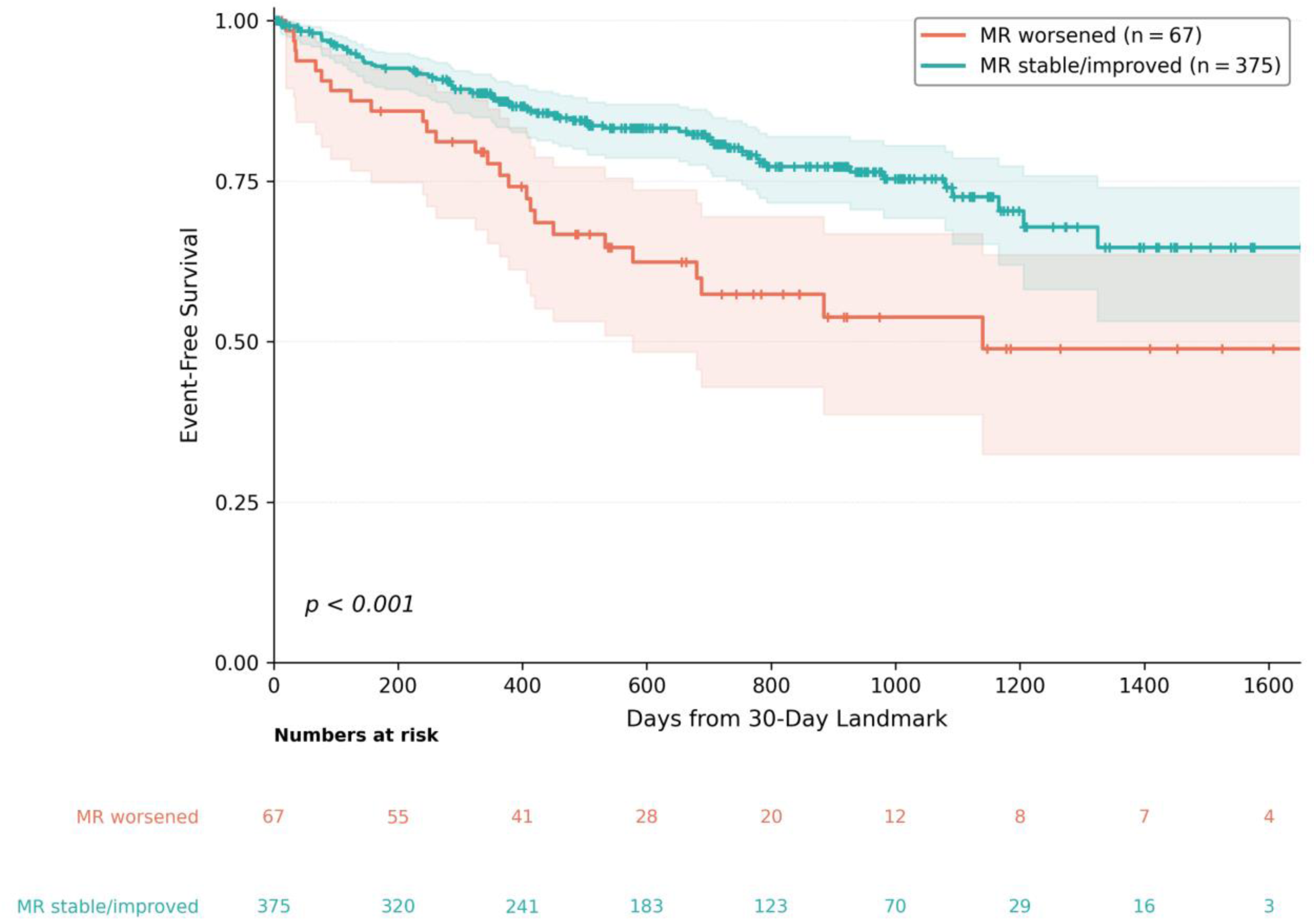
Kaplan-Meier curves for the composite outcome (all-cause death or heart failure hospitalization) by 30-day MR worsening status. Event-free survival after the 30-day landmark is shown for patients with MR worsening versus stable or improved MR at the 30-day echocardiographic assessment. MR worsening was defined as a ≥1-grade increase from baseline. The Kaplan-Meier cohort comprised 442 patients (67 with MR worsening and 375 with stable/improved MR). Note: Table 5 reports N=443 for the 30-day landmark Cox model; 1 additional patient censored at exactly 30 days (contributing zero post-landmark follow up) is excluded from the Kaplan-Meier analysis, yielding N=442. Tick marks indicate censored observations. Log-rank p-value: < 0.001.

**Figure 3.**
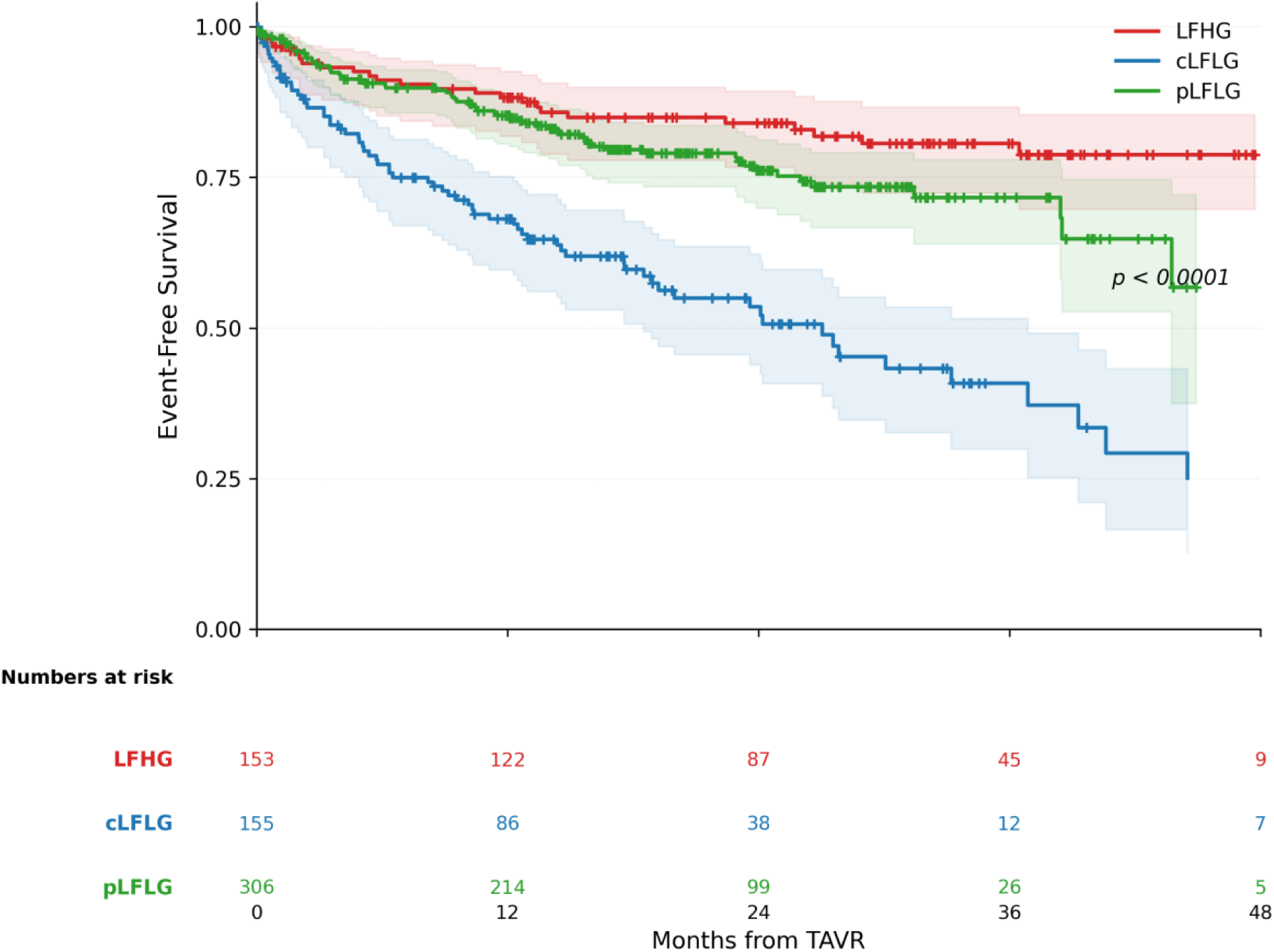
Kaplan-Meier curves for composite all-cause death or heart failure hospitalization by low-flow aortic stenosis phenotype. Event-free survival from the time of TAVR is shown for the full LFAS analytic cohort: LFHG (n=153), cLFLG (n=155), and pLFLG (n=306). Curves are truncated at 48 months for visualization. Tick marks indicate censored observations. Event-free survival differed significantly across LFAS phenotypes by log-rank testing (p < 0.0001), with the lowest event-free survival observed among cLFLG patients.

## Discussion

### Principal Findings

The principal findings of this study are fivefold. First, early MR trajectory after TAVR differed significantly across LFAS phenotypes, with the highest rate of 30-day MR worsening observed in cLFLG. At 30 days, cLFLG patients had nearly a three-fold higher crude rate of MR worsening compared with LFHG patients and nearly five-fold higher adjusted odds of MR worsening in the fully adjusted model. Second, although the unadjusted distribution of MR change at 1 year did not differ significantly across phenotypes, cLFLG remained independently associated with MR worsening in adjusted logistic models at both 30 days and 1 year. Third, MR worsening was associated with worse unadjusted outcomes at 30 days but was not independently associated with the composite endpoint after multivariable adjustment. Fourth, LFAS phenotype, particularly cLFLG, remained the dominant predictor of adverse clinical outcomes. Fifth, a substantial residual MR burden persisted after TAVR, with MR worsening observed in 15.1% of patients at 30 days and 16.9% at 1 year, highlighting an ongoing clinical challenge in this population.

This study provides novel insights into the phenotype-specific evolution of MR following TAVR in patients with LFAS. We demonstrate that MR trajectory varies across LFAS subtypes in a pattern consistent with mortality outcomes across the spectrum of LFAS disease.^14^ Adjusted models identified cLFLG as independently associated with MR worsening. These findings support the importance of phenotype-based risk stratification and surveillance strategies in LFAS patients undergoing TAVR.

### Interpretation of Phenotype-specific MR Evolution

The observed phenotype-specific patterns likely reflect distinct pathophysiological mechanisms underlying each LFAS subtype. In cLFLG, the combination of reduced LVEF and low transvalvular gradient suggests advanced myocardial dysfunction with possible hibernating myocardium.^15^ The high rate of MR worsening in this group may reflect more advanced myocardial phenotype with worse ventricular dysfunction, impaired contractile reserve, and greater comorbidity burden. Unlike paradoxical LFLG-AS where the low-gradient may result from small cavity size, the adverse ventricular remodeling, annular dilation, persistent elevation in filling pressures, or impaired contractile reserve in cLFLG may limit the potential for MR improvement even after afterload reduction.^16^ Because MR mechanism was not available in this study, these mechanistic explanations remain hypothesis-generating.

The persistence of an independent association between cLFLG and MR worsening in adjusted models, despite a nonsignificant unadjusted 1-year trajectory comparison, may reflect the importance of baseline MR severity and competing patient selection effects. Higher baseline MR grade was inversely associated with subsequent worsening, likely because patients with more advanced MR at baseline had less room to worsen on an ordinal grading scale. Adjustment for baseline MR severity may therefore reveal an association between cLFLG phenotype and MR worsening that is less apparent in unadjusted distributional comparisons. In addition, 1-year echocardiographic follow up was limited to a healthier survivor cohort, which may have attenuated unadjusted differences across phenotypes.

### MR Worsening and Clinical Outcomes

Given the limited number of clinical events per subgroup it is challenging to draw significant conclusions beyond MR evolution trends across the entire low-flow cohort. This makes it challenging to predict the specific role of MR evolution in each subtype of LFAS - whether as an innocent bystander or a driver of the worst outcomes among the LFAS spectrum. The high mortality in classical LFLG-AS combined with frequent MR progression raises the possibility of a synergistic relationship, where MR worsening may amplify the already compromised hemodynamics in these patients. Although cLFLG patients had both greater MR instability and worse clinical outcomes, exploratory interaction analyses did not demonstrate a statistically significant phenotype-specific effect of MR worsening on outcomes. This should be interpreted cautiously rather than as evidence of no interaction. Once patients were stratified by LFAS subtype, MR worsening status, landmark timepoint, and clinical outcome, subgroup event counts were limited, particularly in the 1-year cohort. As a result, the interaction analyses were likely underpowered to detect modest phenotype-specific differences in the prognostic impact of MR worsening. Larger studies of subpopulations of LFAS are needed to answer this important question as it may have some bearing on the opportunity for mitral valve intervention in subgroups of LFAS, which has been shown to be useful in certain contexts.^17^

The pLFLG phenotype showed intermediate rates of MR worsening (13.0% at 30 days and 17.6% at 1 year), consistent with a more heterogeneous pathophysiology.^2, 18^ While some patients may experience favorable remodeling from afterload reduction, others may have fixed geometric abnormalities that predispose to MR progression.

LFHG patients with preserved contractility despite low-flow demonstrated the most stable MR trajectory with the lowest worsening rates. This stability may reflect less advanced myocardial disease and more predictable hemodynamic responses to afterload reduction.^19^ The absence of significant prognostic associations in this group suggests that MR evolution is less critical when underlying ventricular function remains preserved.

### Clinical Implications

Our findings have several important clinical implications. First, pre-procedural counseling and patient selection should incorporate phenotype-specific expectations regarding MR evolution.^2, 20^ Patients with cLFLG should be counseled that MR may be less likely to improve after TAVR, and clinicians should recognize that relief of aortic valve obstruction may not fully reverse the mitral-valve burden in this subgroup.^21^ This may influence decisions regarding concomitant or staged mitral intervention in selected cases.

Second, post-TAVR surveillance protocols should be tailored by phenotype. cLFLG patients as well as those with MR worsening warrant closer echocardiographic monitoring, particularly beyond the immediate post-procedural period, as this may identify patients with a more advanced LFAS substrate who require closer follow up and optimization of heart failure therapy. The persistent MR burden observed across all phenotypes (15.1% worsening at 30 days and 16.9% worsening at 1 year) highlights unmet therapeutic needs.^22^ Development of phenotype-specific algorithms for patient selection and timing of mitral intervention could optimize outcomes.

### Comparison with Previous Studies

Our results both confirm and extend previous observations regarding MR evolution after TAVR. The overall rates of improvement we observed (27%) and worsening (19%) align with prior reports, though most studies have not been stratified by AS phenotype.^21, 22, 24^

The lack of independent association between MR change and mortality in unstratified analysis corroborates several recent studies questioning the prognostic significance of MR evolution.^23, 25^ However, our phenotype-specific findings reconcile apparent contradictions in the literature, suggesting that prognostic associations may be obscured when heterogeneous populations are analyzed together.

The particularly high MR volatility in low LVEF patients (our cLFLG group) has been noted previously, though not in the context of comprehensive LFAS phenotyping.^25^ Our findings extend this observation by demonstrating that low-flow status, rather than reduced ejection fraction alone, drives differential MR behavior and prognostic implications.

### Limitations

Several limitations merit consideration. First, the single-center design may limit generalizability, though our quaternary care center’s diverse population enhances external validity.

Second, MR mechanism was not consistently available, limiting our ability to distinguish primary from secondary MR or to determine whether phenotype-specific trajectories differed by MR etiology. Third, MR severity was graded using a six-level clinical scale derived from routine echocardiographic reporting rather than uniform quantitative MR parameters. In this scale, moderate-or-greater MR was defined as grade ≥3. Although this approach reflects real-world clinical interpretation, it differs from formal quantitative MR adjudication and may limit reproducibility. Fourth, echocardiographic follow up was incomplete and subject to survivorship bias, particularly at 1 year. Patients without 1-year echocardiographic follow up had higher mortality and greater comorbidity burden than those with follow up (Supplemental Table S1); therefore, late MR trajectory findings should be interpreted as applying to patients who survived and returned for interval echocardiographic reassessment. Fifth, our sample size, while limiting some subgroup analyses, was sufficient to identify important phenotype-specific differences in MR worsening rates and confirm the dominant prognostic role of cLFLG phenotype. Finally, we could not rigorously assess procedural factors or medical therapy optimization, representing opportunities for future therapeutic interventions.

### Future Directions

Our findings of differential MR worsening rates across LFAS phenotypes establish several actionable research priorities. Risk stratification tools should incorporate both phenotype and MR trajectory. The approximately one quarter of MR worsening rate (26.2% at 30 days and 24.1% at 1 year) in cLFLG patients identifies a particularly high-risk group warranting close surveillance and increasingly optimized heart failure regimens as well as consideration of additional interventions. The absence of statistically significant interaction should not be interpreted as evidence that the prognostic impact of MR worsening is uniform across LFAS phenotypes. Larger multicenter cohorts are needed to determine whether MR trajectory modifies risk differently across subtypes. The clinically meaningful rates of MR worsening (15.1% at 30 days, 16.9% at 1 year) justify continued focus on this problem. Even without proven mortality impact, the symptom burden and heart failure hospitalizations associated with residual MR remain important therapeutic targets. Early identification of the approximate quarter of cLFLG patients who are likely to experience MR worsening represents a key opportunity. Biomarkers, advanced imaging, or clinical predictors could enable preventive strategies. Given that cLFLG patients face both the highest baseline risk and greatest MR instability, trials of upfront combined aortic-mitral intervention in selected patients merit consideration, particularly as transcatheter mitral technologies mature.

## Conclusions

This study provides novel insights into phenotype-specific MR evolution following TAVR in LFAS patients. We demonstrate three key findings: First, early MR trajectory differed significantly by LFAS phenotype, with cLFLG patients experiencing the highest rate of 30-day MR worsening. Although unadjusted 1-year MR-change distributions did not differ significantly across phenotypes, cLFLG remained independently associated with MR worsening in adjusted models at both 30 days and 1 year. Second, LFAS phenotype, particularly cLFLG, remained the strongest determinant of adverse clinical outcomes after TAVR, whereas MR worsening was associated with worse unadjusted outcomes but was not independently associated with the composite endpoint after multivariable adjustment. Third, MR worsening occurred in a substantial minority of LFAS patients after TAVR, underscoring the persistent clinical burden of residual or progressive MR in this population, representing a substantial clinical challenge regardless of statistical associations with mortality.

These findings support phenotype-informed post-TAVR surveillance, particularly for patients with cLFLG, who appear to have both greater MR instability and worse clinical outcomes. The absence of statistically significant phenotype-MR interactions should be interpreted cautiously given limited subgroup event counts, and should not be taken as evidence that the prognostic impact of MR worsening is uniform across LFAS subtypes. Larger multicenter studies are needed to determine whether MR trajectory modifies risk differently across LFAS phenotypes and whether selected patients with persistent or worsening MR may benefit from staged mitral intervention. Our work establishes that both AS phenotype and MR trajectory deserve attention in LFAS management, with phenotype driving baseline risk and MR evolution potentially modifying that risk. Future studies should build on these phenotype-specific insights to develop targeted surveillance and intervention strategies that address the substantial burden of residual MR in this complex population.

## Data Availability

The data underlying this study are not publicly available due to patient privacy and institutional restrictions. De-identified data may be available from the corresponding author upon reasonable request and subject to appropriate institutional approvals.

## Acknowledgements

We thank Yash Prakash, MD, for assistance with early data collection and curation, and for helpful preliminary project discussions.

We also thank members of the Lerakis Cardiovascular Imaging Laboratory for assistance with data collection and refinement of project ideas.

During preparation of this manuscript, the authors used Claude by Anthropic and ChatGPT by OpenAI to assist with statistical code debugging, identification of potentially relevant literature, and review of manuscript for internal consistency. AI assistance was used to support review of the Results, Discussion, tables, figures, captions. The authors independently reviewed, edited, and verified all AI-assisted suggestions including statistical outputs, literature references, and manuscript text, and take full responsibility for the final content of the publication.

## Notes

### Competing Interest Statement

The authors have declared no competing interest.

### Author Declarations

The Institutional Review Board of the Icahn School of Medicine at Mount Sinai gave ethical approval for this work (Project ID: STUDY-22-0036). The requirement for informed consent was waived due to the retrospective design.

