## Supplemental Table S1 for "Mitral regurgitation trajectories after transcatheter aortic valve replacement are phenotype specific across low-flow aortic stenosis subtypes"

### Supplemental Tables

**Supplemental Table S1.** Baseline characteristics and outcomes by MR trajectory analytic cohort availability

| Characteristic | Echo Available | Echo Unavailable | p-value |
|---|---|---|---|
| | *n (%)[1]* | *n (%)[1]* | |
| **Panel A. 30-Day MR Trajectory Availability [Echo available: N = 443 \| Echo unavailable: N = 171]** | | | |
| Age, years, median (IQR) | 82.4 (75.8–87.5) | 83.8 (77.1–88.7) | 0.110 |
| Female sex, n (%) | 279 (63.0%) | 103 (60.2%) | 0.592 |
| LFAS phenotype, n (%) | | | 0.084 |
| LFHG | 121 (27.3%) | 32 (18.7%) | |
| cLFLG | 107 (24.2%) | 48 (28.1%) | |
| pLFLG | 215 (48.5%) | 91 (53.2%) | |
| STS-PROM, %, median (IQR) | 3.3 (2.1–5.1) | 3.7 (2.4–5.8) | 0.065 |
| Baseline MR grade, median (IQR) | 2.0 (1.0–2.0) | 2.0 (2.0–2.0) | **0.009** |
| Moderate-or-greater MR (grade ≥3), n (%) | 65 (14.7%) | 34 (19.9%) | 0.147 |
| Chronic kidney disease, n (%) | 143 (32.3%) | 70 (40.9%) | 0.054 |
| Atrial fibrillation, n (%) | 163 (36.8%) | 61 (35.7%) | 0.869 |
| Coronary artery disease, n (%) | 212 (47.9%) | 96 (56.1%) | 0.080 |
| Prosthesis–patient mismatch, n/N (%) | 51/303 (16.8%) | 3/30 (10.0%) | 0.441 |
| **Clinical Outcomes** | | | |
| All-cause mortality, n (%) | 69 (15.6%) | 48 (28.1%) | **<0.001** |
| Heart failure hospitalization, n (%) | 41 (9.3%) | 25 (14.6%) | 0.075 |
| Composite death or HFH, n (%) | 98 (22.1%) | 64 (37.4%) | **<0.001** |
| **Panel B. 1-Year MR Trajectory Availability [Echo available: N = 290 \| Echo unavailable: N = 324]** | | | |
| Age, years, median (IQR) | 82.0 (76.2–87.2) | 83.5 (76.5–88.8) | 0.112 |
| Female sex, n (%) | 175 (60.3%) | 207 (63.9%) | 0.412 |
| LFAS phenotype, n (%) | | | **0.002** |
| LFHG | 77 (26.6%) | 76 (23.5%) | |
| cLFLG | 54 (18.6%) | 101 (31.2%) | |
| pLFLG | 159 (54.8%) | 147 (45.4%) | |
| STS-PROM, %, median (IQR) | 3.0 (1.9–4.8) | 3.8 (2.5–5.7) | **<0.001** |
| Baseline MR grade, median (IQR) | 2.0 (1.0–2.0) | 2.0 (1.0–2.0) | 0.063 |
| Moderate-or-greater MR (grade ≥3), n (%) | 36 (12.4%) | 63 (19.4%) | **0.024** |
| Chronic kidney disease, n (%) | 83 (28.6%) | 130 (40.1%) | **0.004** |
| Atrial fibrillation, n (%) | 89 (30.7%) | 135 (41.7%) | **0.006** |
| Coronary artery disease, n (%) | 137 (47.2%) | 171 (52.8%) | 0.197 |
| Prosthesis–patient mismatch, n/N (%) | 26/169 (15.4%) | 28/164 (17.1%) | 0.788 |
| **Clinical Outcomes** | | | |
| All-cause mortality, n (%) | 22 (7.6%) | 95 (29.3%) | **<0.001** |
| Heart failure hospitalization, n (%) | 16 (5.5%) | 50 (15.4%) | **<0.001** |
| Composite death or HFH, n (%) | 36 (12.4%) | 126 (38.9%) | **<0.001** |

[1] Unless otherwise noted, categorical variables are shown as n (%) with denominator equal to the number of patients with non-missing values for that variable within each group (consistent with the p-value denominator). Continuous variables are shown as median (IQR). For PPM, denominator reflects patients with PPM data available.

Clinical outcomes are shown descriptively to illustrate landmark/survivorship selection and were not used to define baseline comparability.

**Statistical tests:** Wilcoxon rank-sum test for continuous variables; chi-square or Fisher exact test (when any expected cell count < 5) for categorical variables. Bold p-values indicate $p < 0.05$.

**Echo availability definitions:** Groups correspond to the MR trajectory analytic cohorts used in the primary manuscript analyses. 30-day MR trajectory available: echocardiogram within 15– 90 days post-TAVR with computable MR change (strict pre-event rule applied: echoes occurring after a composite endpoint event were excluded). 1-year MR trajectory available: echocardiogram within 185–545 days post-TAVR with computable MR change, additionally requiring that no composite endpoint event occurred within 365 days (landmark filter). Patients without a qualifying echocardiogram, or with a pre-landmark composite event, are included in the 'unavailable' group.

**Abbreviations:** AF, atrial fibrillation; CAD, coronary artery disease; CKD, chronic kidney disease; HFH, heart failure hospitalization; IQR, interquartile range; LFAS, low-flow aortic stenosis; LFHG, low-flow high-gradient; cLFLG, classical low-flow low-gradient; MR, mitral regurgitation; pLFLG, paradoxical low-flow low-gradient; PPM, prosthesis–patient mismatch; STS-PROM, Society of Thoracic Surgeons Predicted Risk of Mortality; TAVR, transcatheter aortic valve replacement.
